# Efficacy of two doses of COVID-19 vaccine against severe COVID-19 in those with risk conditions and residual risk to the clinically extremely vulnerable: the REACT-SCOT case-control study

**DOI:** 10.1101/2021.09.13.21262360

**Authors:** Paul M McKeigue, David A McAllister, Chris Robertson, Sharon Hutchinson, Stuart McGurnaghan, Diane Stockton, Helen M Colhoun, for the PHS COVID-19 Epidemiology and Research Cell

**Affiliations:** Usher Institute, College of Medicine and Veterinary Medicine, University of Edinburgh, Teviot Place, Edinburgh EH8 9AG, Scotland; Institute of Health and Wellbeing, University of Glasgow, 1 Lilybank Gardens, Glasgow G12 8RZ; Department of Mathematics and Statistics, University of Strathclyde, 16 Richmond Street, Glasgow G1 1XQ; School of Health and Life Sciences, Glasgow Caledonian University. SH - Professor of Epidemiology and Population Health; Institute of Genetics and Cancer, College of Medicine and Veterinary Medicine, University of Edinburgh, Western General Hospital Campus, Crewe Road, Edinburgh EH4 2XUC, Scotland; Public Health Scotland, Meridian Court, 5 Cadogan Street, Glasgow G2 6QE

## Abstract

**Objectives:** To determine whether COVID-19 efficacy varies with clinical risk category and to investigate risk factors for severe COVID-19 in those who have received two doses of vaccine.

**Design:** Matched case-control study (REACT-SCOT).

**Setting:** Population of Scotland from 1 December 2020 to 8 September 2021.

**Main outcome measure:** Severe COVID-19, defined as cases with entry to critical care or fatal outcome.

**Results:** Efficacy against severe COVID-19 of two doses of vaccine was 94% (95 percent CI 93% to 96%) in those without designated risk conditions, 89% (95 percent CI 86% to 91%) in those with moderate risk conditions, but only 73% (95 percent CI 64% to 79%) in those designated as clinically extremely vulnerable (CEV) and eligible for shielding. Of the 641 cases of severe COVID-19 in double-vaccinated individuals, 47% had moderate risk conditions and 38% were CEV. In the double-vaccinated CEV group, the rate ratio for severe disease (with no risk condition as reference category) was highest in solid organ transplants at 101 (95% CI 47 to 214) but even in this subgroup the absolute risk of severe COVID-19 was low (35 cases in 23678 person-months of follow-up).

**Conclusions:** Two doses of vaccine protect against severe COVID-19 in CEV individuals but the residual risk in double-vaccinated individuals remains far higher in those who are CEV than in those who are not. These results lay a basis for determining eligibility for additional measures including passive immunization to protect those at highest risk.

## Introduction

The REACT-SCOT matched case-control study was established by Public Health Scotland at the beginning of the epidemic to investigate risk factors for severe COVID-19 [1]. Using this framework, we have reported on the relation of severe COVID-19 to risk conditions including those designated as clinically extremely vulnerable (CEV) and therefore eligible for the shielding programme in Scotland [1–3]. We have previously reported a comparison of the efficacy of vaccination against severe COVID-19 between CEV people and those with moderate risk conditions or no risk conditions, based on data up to 16 March 2021 [4]. At this time few individuals had received a second dose of vaccine, but since then most CEV individuals have received two doses. This report updates and extends the earlier analyses, with the following objectives:

1. To determine whether vaccine efficacy varies with risk category and CEV status now that more data, including exposure to two doses of vaccine, are available and now that the Delta variant is the dominant variant in Scotland.
2. To investigate risk factors for severe or hospitalised COVID-19 in those who have received two doses of vaccine.

## Methods

We used the REACT-SCOT study to take advantage of data linkages already established. The design has been described in detail previously [1]. In brief, for every incident case of COVID-19 in the population ten controls matched for one-year age, sex and primary care practice and alive on the day of presentation of the case that they were matched to were selected using the Community Health Index database. COVID-19 cases are those with a positive nucleic acid test, or a hospital admission or death with COVID-19 ICD-10 codes. The REACT-SCOT case-control dataset is refreshed regularly and is linked to the vaccination database and to the regularly updated dataset of all individuals deemed eligible for the shielding programme. Though the data extract included cases presenting up to 22 September 2021, the analyses reported here are restricted to cases and controls presenting from 1 December 2020 to 8 September 2021, ensuring follow-up for at least 14 days after presentation date to allow cases to be classified as severe or hospitalised.

### Classification of risk categories

As previously [1], to minimise ascertainment bias we pre-specified the primary outcome measure as severe COVID-19, defined as diagnosed cases with entry to critical care within 28 days of presentation or fatal outcome (any death within 28 days of a positive test or any death for which COVID-19 was coded as underlying cause). Cases and controls were classified into three broad risk categories: no risk condition; at least one of the moderate risk conditions designated by public health agencies [1]; or CEV (eligible for shielding) [3]. For further analyses, the CEV category was subdivided as described previously into six categories: solid organ transplant, specific cancers, severe respiratory conditions, other rare conditions, on immunosuppressants, and additional conditions [3]. This corresponds to the list used by Public Health Scotland [5], after combining the small numbers in the group “pregnant with heart disease with the”other conditions” category. For additional analyses the category “specific cancers” was split to allocate cancers of blood-forming organs (ICD-10 codes C81-C88, C90-C96) to a separate category.

### Statistical analysis

The matched design controls for age, sex, general practice and calendar time to single day. Covariates included in the models were those that have been previously identified as strong predictors of severe disease in this population: care home residence, number of adults in household, number of non-cardiovascular drug classes dispensed and recent hospital stay [1–3]. For care home residents the number of of adults in the household was coded as 1 to ensure that these two variables are not confounded. The number of non-cardiovascular drug classes was calculated as the number of distinct BNF subparagraph codes for which a prescription was dispensed between 15 and 255 days before presentation date. The number of hospital diagnoses was calculated as the number of distinct ICD-10 chapters represented at least once in hospital discharge records between 25 days and 5 years before presentation date. Recent hospital stay was defined as any in-patient stay from 5 to 14 days before presentation date.

Vaccination status was coded as the number of doses administered at least 14 days before presentation date. Vaccine doses administered less than 14 days before presentation date were ignored. The effect of vaccination in each of the clinical vulnerability categories was estimated in a conditional logistic regression model specifying effects *β*_*R*2_, …, *β*_*RJ*_ for the log rate ratio associated with risk categories 2 to *J* (*β*_*R*1_ = 0 for the reference category *J* = 1), and nested effects *β*_*V* 1_, …, *β*_*V J*_ for the log rate ratio associated with vaccination in each of the *J* risk categories. With this incidence density sampling design, the conditional odds ratio is the rate ratio. The efficacy of vaccination is 1 minus the rate ratio [6]. The unconditional odds ratios calculated from frequency tables of the vaccination status of cases and controls in each risk group cannot be used to estimate rate ratios [7,8].

### Cohort analysis of the shielding list

The case-control study estimates only rate ratios. To investigate how the absolute rates of severe disease in those listed as clinically extremely vulnerable have changed with the vaccination programme rollout, we also undertook a cohort analysis of all individuals who had ever been on the shielding list. A Poisson regression model was fitted to the cohort formatted with one observation per 28-day person-time interval, and individuals censored at first diagnosis of COVID-19. Event status was defined as severe COVID-19 presenting within the person-time interval, and the covariates were sex, baseline age, and shielding eligibility group. The baseline hazard rate was modelled as a natural spline function of calendar time with 6 degrees of freedom. To allow comparison with how the rates of all diagnosed cases varied with calendar time, a similar model was fitted with event status defined as any diagnosed case.

## Results

### Relation of vaccine efficacy to risk conditions

As shown in Table 1 from 1 December 2020 to 8 September 2021 there were 5644 cases of severe COVID-19 in the total population of Scotland of whom 28% had no designated risk condition, 51% had a moderate risk condition and 21% were CEV. The distributions in cases and controls of other risk factors included as covariates in the models are shown for completeness. Overall 80% of severe cases arising in this period were not yet vaccinated. Table S1 shows the same tabulation with case definition broadened to include all 21671 hospitalised cases and their matched controls.

**Table 1.**
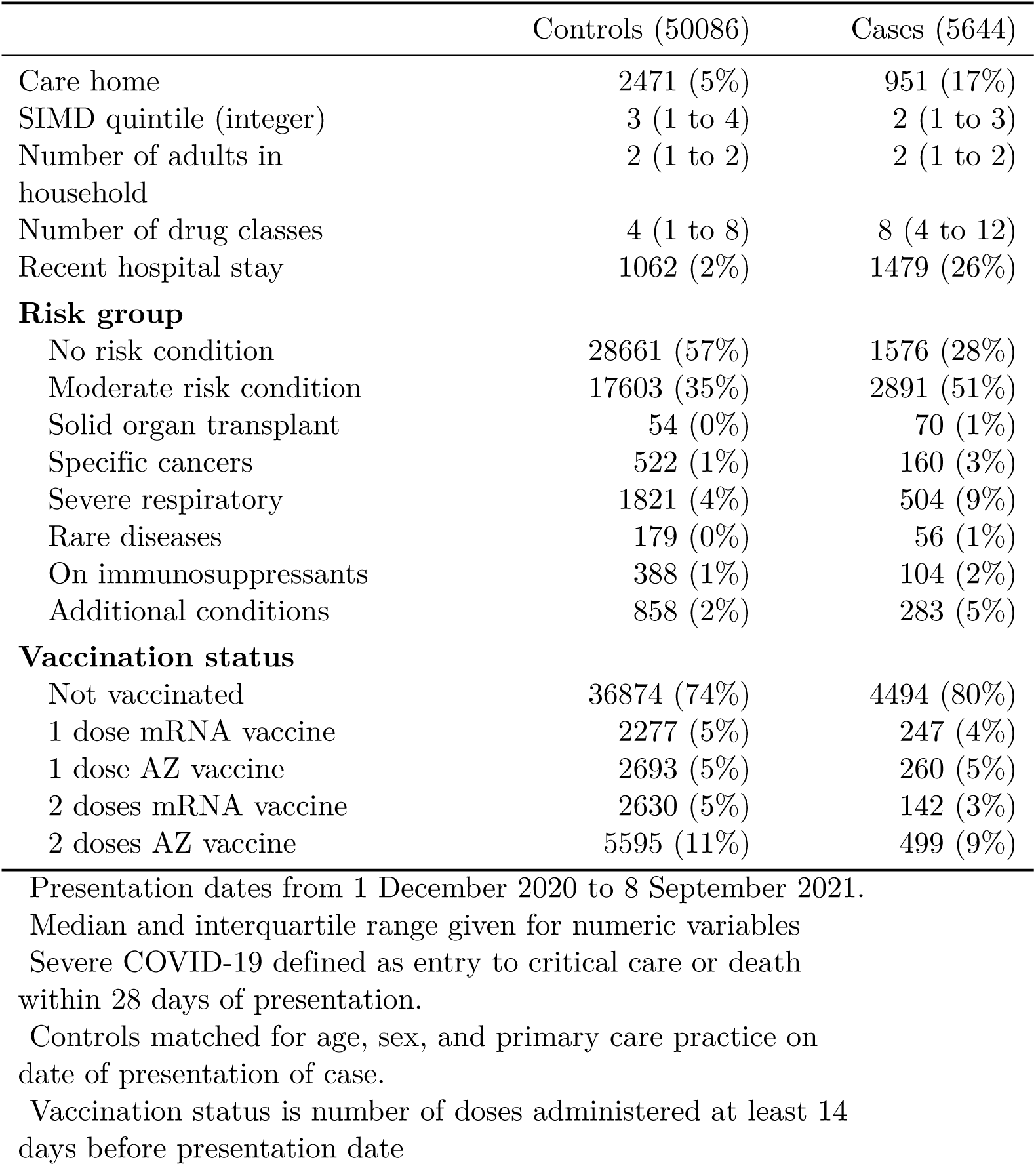
Numbers of cases of severe COVID-19 and matched controls, by risk group and vaccine product

Table 2 shows rate ratios associated with 1 and 2 vaccine doses (with 0 doses as reference category) in each of the three broad risk categories from the conditional logistic regression model. The model included risk categories (no risk conditions as reference category), care home residence, number of adults in household, number of drug classes and recent hospital stay as covariates. The rate ratio associated with two doses of vaccine was 0.06 (95% CI 0.04 to 0.07) in those without risk conditions, 0.11 (95% CI 0.09 to 0.14) in those with moderate risk conditions, and 0.27 (95% CI 0.21 to 0.36) in the CEV group. On the scale of efficacy, these estimates are equivalent to 94% (95 percent CI 93% to 96%) in those without risk conditions, 89% (95 percent CI 86% to 91%) in those with moderate risk conditions, and 73% (95 percent CI 64% to 79%) in the CEV group. Table S2 shows the same model with case definition broadened to include all hospitalised cases and their matched controls: the efficacy of 2 doses against hospitalisation also was lower at 68% (95 percent CI 63% to 71%) in the CEV than in the other two risk categories. The difference in vaccine efficacy against hospitalisation between the CEV group and the other two risk categories was not explained by differences in time since second dose.

**Table 2.**
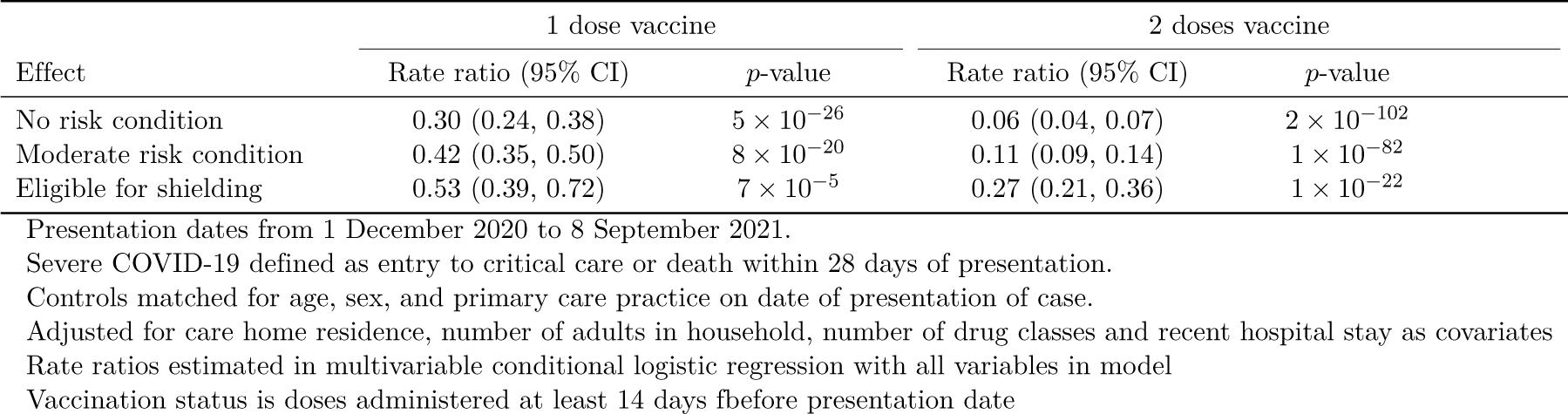
Rate ratios for severe COVID-19 within risk groups associated with vaccine dose: unvaccinated as reference category

| Effect | 1 dose vaccine |  | 2 doses vaccine |  |
| --- | --- | --- | --- | --- |
|  | Rate ratio (95% CI) | *p*-value | Rate ratio (95% CI) | *p*-value |
| No risk condition | 0.30 (0.24, 0.38) | $5 \times 10^{-26}$ | 0.06 (0.04, 0.07) | $2 \times 10^{-102}$ |
| Moderate risk condition | 0.42 (0.35, 0.50) | $8 \times 10^{-20}$ | 0.11 (0.09, 0.14) | $1 \times 10^{-82}$ |
| Eligible for shielding | 0.53 (0.39, 0.72) | $7 \times 10^{-5}$ | 0.27 (0.21, 0.36) | $1 \times 10^{-22}$ |

Presentation dates from 1 December 2020 to 8 September 2021.
Severe COVID-19 defined as entry to critical care or death within 28 days of presentation.
Controls matched for age, sex, and primary care practice on date of presentation of case.
Adjusted for care home residence, number of adults in household, number of drug classes and recent hospital stay as covariates
Rate ratios estimated in multivariable conditional logistic regression with all variables in model
Vaccination status is doses administered at least 14 days before presentation date

Table 3 shows rate ratios associated with vaccination by risk group with the CEV group subdivided into six categories. Rate ratios were higher (and thus efficacy was lower) in all CEV subgroups than in those without risk conditions or with moderate risk conditions, but the confidence intervals were too wide for comparisons of efficacy between CEV subgroups. Within the “Specific cancers” group, efficacy of two doses did not differ between those with blood cancers and those with other types of cancer but the confidence intervals were wide. Table S3 shows rate ratios for the first and second dose with the case definition broadened to include all hospitalised cases and their matched controls. In this table the highest rate ratio (lowest efficacy) is in solid organ transplant recipients but again the confidence intervals are too wide for comparisons of efficacy between CEV subgroups.

**Table 3.**
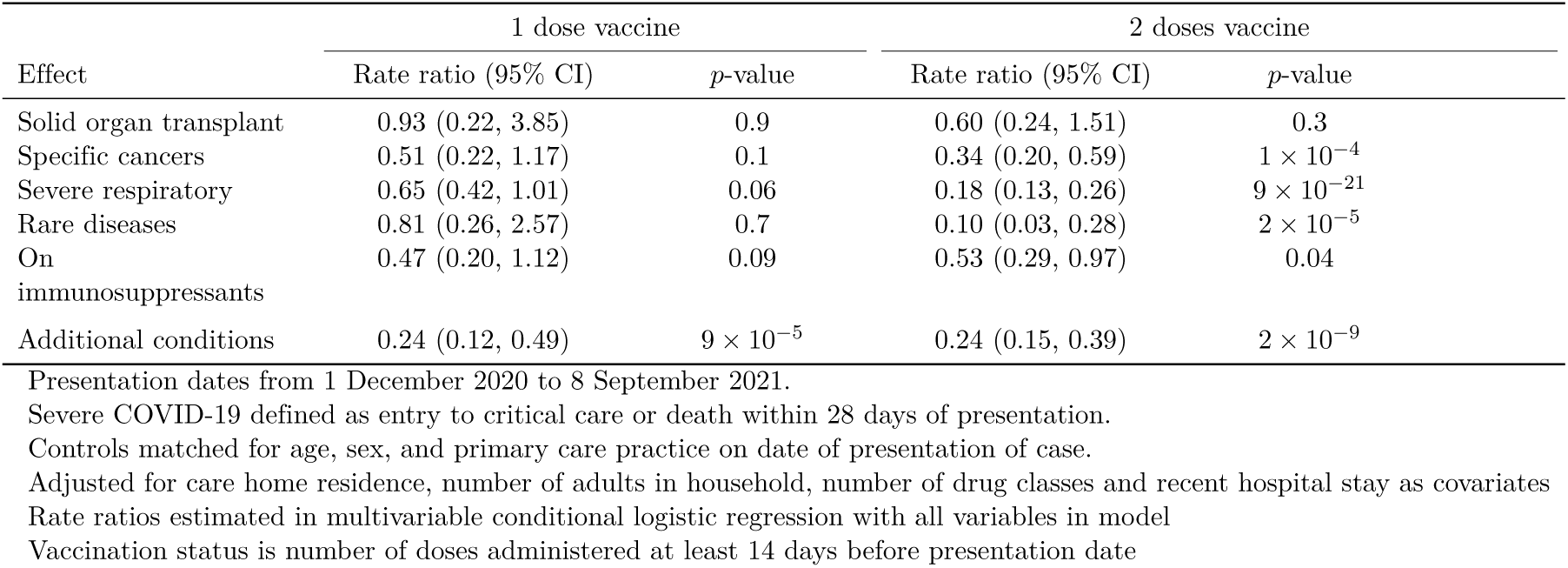
Rate ratios for severe COVID-19 associated with vaccine dose within clinically extremely vulnerable subgroups/2, 1

| Effect | 1 dose vaccine |  | 2 doses vaccine |  |
| --- | --- | --- | --- | --- |
|  | Rate ratio (95% CI) | *p*-value | Rate ratio (95% CI) | *p*-value |
| Solid organ transplant | 0.93 (0.22, 3.85) | 0.9 | 0.60 (0.24, 1.51) | 0.3 |
| Specific cancers | 0.51 (0.22, 1.17) | 0.1 | 0.34 (0.20, 0.59) | $1 \times 10^{-4}$ |
| Severe respiratory | 0.65 (0.42, 1.01) | 0.06 | 0.18 (0.13, 0.26) | $9 \times 10^{-21}$ |
| Rare diseases | 0.81 (0.26, 2.57) | 0.7 | 0.10 (0.03, 0.28) | $2 \times 10^{-5}$ |
| On immunosuppressants | 0.47 (0.20, 1.12) | 0.09 | 0.53 (0.29, 0.97) | 0.04 |
| Additional conditions | 0.24 (0.12, 0.49) | $9 \times 10^{-5}$ | 0.24 (0.15, 0.39) | $2 \times 10^{-9}$ |

Presentation dates from 1 December 2020 to 8 September 2021.
Severe COVID-19 defined as entry to critical care or death within 28 days of presentation.
Controls matched for age, sex, and primary care practice on date of presentation of case.
Adjusted for care home residence, number of adults in household, number of drug classes and recent hospital stay as covariates
Rate ratios estimated in multivariable conditional logistic regression with all variables in model
Vaccination status is number of doses administered at least 14 days before presentation date

We examined whether there was any difference in efficacy by class of vaccine and whether any such differences varied by risk category. Table 4 shows rate ratios associated with 1 and 2 doses of each class of vaccine (with 0 doses as reference category) in each of the three broad risk categories. Table S4 shows the same model with case definition broadened to include all hospitalised cases and their matched controls. Although the confidence intervals for the rate ratios associated with 1 or 2 doses of each vaccine product in the CEV group are wide, it is clear that for both classes of vaccine product the efficacy of 2 doses is lower in the CEV group than in those with moderate risk conditions or no risk conditions. Efficacy of two doses against severe COVID-19 in the CEV group did not differ between the AstraZeneca vaccine [72% (95 percent CI 63% to 79%)] and the mRNA vaccines [73% (95 percent CI 59% to 83%)] Further comparison of efficacy of the two vaccine types in CEV individuals has been reported elsewhere [9].

**Table 4.** Rate ratios for severe COVID-19 within risk groups: vaccine dose and product encoded as categoric variable with unvaccinated as reference category

| Effect | 1 dose vaccine |  | 2 doses vaccine |  |
| --- | --- | --- | --- | --- |
|  | Rate ratio (95% CI) | *p*-value | Rate ratio (95% CI) | *p*-value |
| No risk condition: mRNA vaccine | 0.25 (0.18, 0.34) | $7 \times 10^{-18}$ | 0.04 (0.03, 0.06) | $9 \times 10^{-46}$ |
| No risk condition: AZ vaccine | 0.35 (0.26, 0.47) | $3 \times 10^{-12}$ | 0.07 (0.05, 0.09) | $7 \times 10^{-73}$ |
| Moderate risk condition: mRNA vaccine | 0.52 (0.42, 0.65) | $2 \times 10^{-8}$ | 0.07 (0.05, 0.09) | $7 \times 10^{-55}$ |
| Moderate risk condition: AZ vaccine | 0.30 (0.23, 0.40) | $9 \times 10^{-17}$ | 0.13 (0.10, 0.17) | $2 \times 10^{-60}$ |
| Eligible for shielding: mRNA vaccine | 0.48 (0.28, 0.82) | 0.007 | 0.27 (0.17, 0.41) | $3 \times 10^{-9}$ |
| Eligible for shielding: AZ vaccine | 0.52 (0.36, 0.75) | $5 \times 10^{-4}$ | 0.28 (0.21, 0.37) | $3 \times 10^{-19}$ |

Presentation dates from 1 December 2020 to 8 September 2021.
Adjusted for care home residence, number of adults in household, number of drug classes dispensed, recent hospital stay and risk category.
Controls matched for age, sex, and primary care practice on date of presentation of case.
Vaccination status is number of doses administered at least 14 days before presentation date

### Risk factors for severe COVID-19 in the double-vaccinated

Table 5 shows risk factors for severe COVID-19 in cases and controls who had received 2 doses of vaccine at least 14 days before presentation date. Table S5 shows the same analysis with case definition broadened to include hospitalized cases and matched controls who had received 2 doses of vaccine. Only 15% of double-vaccinated severe cases had no designated risk condition: 47% had at least one moderate risk condition and 38% were CEV. Of the variables that we have previously reported as risk factors for severe COVID-19 in the general population, care home residence and socioeconomic deprivation were not associated with severe disease in the double-vaccinated. In this group the risk factors for severe disease were designated risk conditions, other indicators of co-morbidity including numbers of hospital diagnoses and drug classes prescribed and transmission-related factors including number of adults in household and recent hospital stay. A risk score for hospitalised or fatal COVID-19 in the double-vaccinated calculated from the multivariable conditional logistic regression model in Table S5 was able to distinguish severe cases from noncases with a C-statistic of 0.79. Among the CEV groups, the highest rate ratio for severe disease was solid organ transplant recipients: with no risk conditions as reference category, the unadjusted rate ratio associated with receipt of a solid organ transplant among the double-vaccinated was 101 (95% CI 47 to 214) for severe disease and 29.9 (95% CI 21.0 to 42.7) for hospitalisation.

**Table 5.** Risk factors for severe COVID-19 in those who had received 2 doses of vaccine at least 14 days before

|  | Controls<br>(8225) | Cases (641) | Univariate |  | Multivariable |  |
| --- | --- | --- | --- | --- | --- | --- |
|  |  |  | Rate ratio (95%<br>CI) | *p*-value | Rate ratio (95%<br>CI) | *p*-value |
| Care home | 207 (3%) | 57 (9%) | 3.11 (2.14, 4.52) | $2 \times 10^{-9}$ | 1.70 (1.10, 2.63) | 0.02 |
| SIMD quintile (integer) | 3 (1 to 4) | 2 (1 to 4) | 0.79 (0.74, 0.85) | $4 \times 10^{-10}$ | 0.92 (0.84, 1.00) | 0.06 |
| Number of adults in household | 2 (1 to 2) | 2 (1 to 2) | 0.95 (0.88, 1.03) | 0.3 | 1.01 (0.92, 1.10) | 0.9 |
| Number of hospital diagnoses | 0 (0 to 3) | 5 (1 to 8) | 1.28 (1.25, 1.31) | $8 \times 10^{-75}$ | 1.06 (1.02, 1.09) | 0.001 |
| Number of non-cardiovascular drug classes | 2 (1 to 5) | 7 (4 to 10) | 1.27 (1.24, 1.30) | $7 \times 10^{-92}$ | 1.13 (1.10, 1.17) | $6 \times 10^{-18}$ |
| Recent hospital stay | 151 (2%) | 90 (14%) | 7.5 (5.5, 10.2) | $1 \times 10^{-36}$ | 3.83 (2.68, 5.47) | $2 \times 10^{-13}$ |
| **Risk group** |  |  |  |  |  |  |
| No risk condition | 4940 (60%) | 95 (15%) | . | . | . | . |
| Moderate risk condition | 2673 (32%) | 303 (47%) | 5.4 (4.2, 7.0) | $1 \times 10^{-37}$ | 2.93 (2.20, 3.89) | $1 \times 10^{-13}$ |
| Solid organ transplant | 15 (0%) | 35 (5%) | 101 (47, 214) | $7 \times 10^{-33}$ | 31.4 (13.8, 71.2) | $2 \times 10^{-16}$ |
| Specific cancers | 77 (1%) | 36 (6%) | 26.0 (15.8, 43.0) | $3 \times 10^{-37}$ | 12.7 (7.3, 22.0) | $2 \times 10^{-19}$ |
| Severe respiratory diseases | 289 (4%) | 82 (13%) | 13.2 (9.3, 18.7) | $2 \times 10^{-47}$ | 4.75 (3.17, 7.11) | $4 \times 10^{-14}$ |
| Rare diseases | 32 (0%) | 6 (1%) | 8.2 (3.2, 21.1) | $1 \times 10^{-5}$ | 3.24 (1.21, 8.66) | 0.02 |
| On immunosuppressants | 61 (1%) | 31 (5%) | 26.2 (15.1, 45.3) | $2 \times 10^{-31}$ | 13.3 (7.3, 24.3) | $3 \times 10^{-17}$ |
| Additional conditions | 138 (2%) | 53 (8%) | 15.5 (10.2, 23.6) | $6 \times 10^{-38}$ | 6.3 (3.9, 10.0) | $3 \times 10^{-14}$ |

Severe COVID-19 is defined by entry to critical care or fatal outcome
Percentages are column percentages for each variable
Rate ratios are from conditional logistic regression models matched for age, sex and general practice
Univariate rate ratios are for models with a single covariate
Multivariable rate ratios are for a model including all covariates shown in the table

### Shielding cohort

By the date of the latest extract of the shielding list in July 2021, 8230 (4%) of the 202510 individuals who had ever been on the shielding list had been diagnosed with COVID-19, and 18893 (9%) had been removed from the list for other reasons. Fig 1 shows the fitted incidence per month of any diagnosis and of severe cases in Poisson regression models fitted to the entire cohort. The incidence of any diagnosed COVID-19 in the shielding cohort fell from 1 December to a nadir in late April. Despite the incidence of any diagnosed COVID-19 rising steeply from 1 May onwards, the incidence of severe COVID-19 rose only slightly. Table S6 shows the incidence of severe COVID-19 in double-vaccinated CEV individuals, by subgroup. In the highest risk group – solid organ transplant recipients – the incidence was 1.5 per 1000 per month (35 cases in 23678 person-months of follow-up). Of the 35 severe cases among double-vaccinated solid organ transplants, only one had received a transplant less than one year before presentation date.

**Fig 1.**
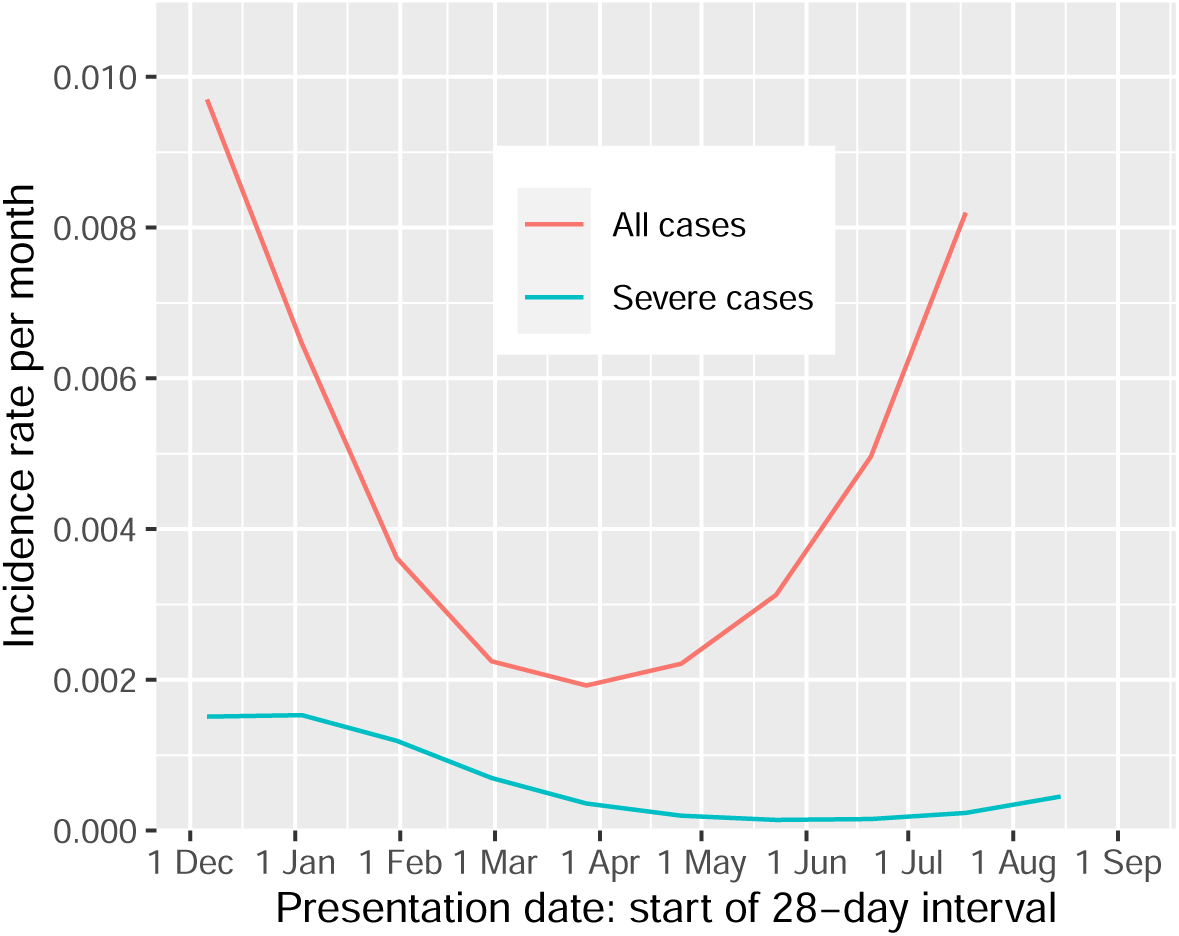
Fitted values for monthly incidence of any case and severe case in the shielding cohort, based on Poisson regression with spline terms for calendar time

## Discussion

### Statement of principal findings

1. Although the efficacy of two doses of vaccine against severe COVID-19 in those without risk conditions remains around 90%, it is now clear that efficacy in clinically extremely vulnerable individuals is somewhat lower at 73%. On the scale of absolute risk reduction, a reduction by three-quarters of fivefold elevated risk in unvaccinated CEV individuals is of course greater than a reduction of baseline risk by nine-tenths.
2. Among double-vaccinated individuals, those who have designated risk conditions or are CEV account for 85% of severe cases and 75% of hospitalised cases.
3. In comparison with double-vaccinated individuals of the same age and sex without risk conditions, double-vaccinated CEV individuals remain at relatively high risk of severe COVID-19. For double-vaccinated solid organ transplant recipients the rate of severe disease is 100-fold higher, though the absolute risk is about 1 in 1000 per month.

### Strengths and limitations

Strengths of this study are the elimination of calendar time effects by the matched case-control design, the comprehensive linkage to e-health records and the focus on severe cases as main outcome measure. Hospitalised cases may include some test-positive individuals whose admission or continued stay in hospital was for another underlying diagnosis: this in turn may lead to underestimation of vaccine efficacy against COVID-19, especially in risk groups with comorbidities. Such misclassification is less likely to occur with the narrow definition of severe COVID-19 used in the REACT-SCOT study.

Even in this large cohort, the numbers of severe cases in some CEV subgroups are too small for vaccine efficacy against severe disease to be estimated accurately within these subgroups. Another limitation is that as most immunosuppressants and drugs for cancer are prescribed only in hospital where prescribing records are not held in electronic form, we cannot study the relation of risk to different classes of immunosuppressant drugs.

### Relation to other studies

Two studies in England have examined vaccine efficacy in the CEV. One estimated that efficacy of the Pfizer vaccine against hospitalisation waned more rapidly in the CEV than in those who were not CEV [10]. The other study estimated efficacy of two doses of any vaccine against symptomatic COVID-19 as 87% (based on combining the age-stratified estimates) in those who had been advised to shield [11]. However without regular scheduled testing, estimation of vaccine efficacy against infection (rather than hospitalisation) is subject to ascertainment bias, and this problem is not entirely overcome by restricting to symptomatic cases or by test-negative case-control designs.

### Policy implications

Our results show that the risk of severe COVID-19 after two doses of vaccine is highest in solid organ transplant recipients, and that this elevated risk is not restricted to the first six months after transplant as the UK Joint Committee on Vaccination and Immunisation apparently assumed in its original recommendation that “a third primary dose be offered to individuals aged 12 years and over with severe immunosuppression” [12]. Subsequent recommendations have broadened the criteria for third primary doses and booster doses. For solid organ transplant recipients, passive immunization therapies may be an option now that they have been licensed in the UK for prevention in “those who have a medical condition making them unlikely to respond to or be protected by vaccination” [13].

## Data Availability

The component datasets used here are available via the Public Benefits and Privacy Panel for Health and Social Care at https://www.informationgovernance.scot.nhs.uk/pbpphsc/ for researchers who meet the criteria for access to confidential data. All source code used for derivation of variables, statistical analysis and generation of this manuscript is available on https://github.com/pmckeigue/covid-scotland_public.

## Declarations

### Public and Patient Involvement statement

This study was conducted under approvals from the Public Benefit and Privacy Panel for Health and Social Care which includes public and patient representatives.

### Ethics approval

This study was performed within Public Health Scotland as part of its statutory duty to monitor and investigate public health problems. Under the UK Policy Framework for Health and Social Care Research set out by the NHS Health Research Authority, this does not fall within the definition of research and ethical review was therefore not required. Individual consent is not required for Public Health Scotland staff to process personal data to perform specific tasks in the public interest that fall within its statutory role. The statutory basis for this is set out in Public Health Scotland’s privacy notice.

### Transparency declaration

PM as the manuscript’s guarantor affirms: that the manuscript is an honest, accurate, and transparent account of the study being reported; that no important aspects of the study have been omitted; and that any discrepancies from the study as originally planned and registered have been explained. This manuscript has been generated directly from the source data by a reproducible research pipeline.

### Funding

No specific funding was received for this study.

### Competing interest

All authors have completed and submitted the ICMJE Form for Disclosure of Potential Conflicts of Interest.

## Acknowledgements

We thank Jen Bishop, Ciara Gribben, Bob Taylor and David Caldwell for undertaking the linkage analysis within Public Health Scotland,

## Supplementary Material

**Table S1.** Numbers of cases of hospitalized or fatal COVID-19 and matched controls, by risk group and vaccine product

|  | Controls (202071) | Cases (21671) |
| --- | --- | --- |
| Care home | 5790 (3%) | 2053 (9%) |
| SIMD quintile (integer) | 3 (1 to 4) | 2 (1 to 3) |
| Number of adults in household | 2 (1 to 2) | 2 (1 to 2) |
| Number of drug classes | 3 (0 to 6) | 6 (2 to 10) |
| Recent hospital stay | 3304 (2%) | 5333 (25%) |
| **Risk group** |  |  |
| No risk condition | 133911 (66%) | 8802 (41%) |
| Moderate risk condition | 55806 (28%) | 9214 (43%) |
| Solid organ transplant | 255 (0%) | 192 (1%) |
| Specific cancers | 1600 (1%) | 506 (2%) |
| Severe respiratory | 5630 (3%) | 1538 (7%) |
| Rare diseases | 575 (0%) | 157 (1%) |
| On immunosuppressants | 1462 (1%) | 330 (2%) |
| Additional conditions | 2832 (1%) | 932 (4%) |
| **Vaccination status** |  |  |
| Not vaccinated | 128417 (64%) | 15369 (71%) |
| 1 dose mRNA vaccine | 11812 (6%) | 788 (4%) |
| 1 dose AZ vaccine | 12562 (6%) | 1440 (7%) |
| 2 doses mRNA vaccine | 16983 (8%) | 774 (4%) |
| 2 doses AZ vaccine | 32213 (16%) | 3294 (15%) |

Presentation dates from 1 December 2020 to 8 September 2021.
Median and interquartile range given for numeric variables
Controls matched for age, sex, and primary care practice on date of presentation of case.
Vaccination status is number of doses administered at least 14 days before presentation date

**Table S2.** Rate ratios for hopitalised or fatal COVID-19 within risk groups associated with vaccine dose: unvaccinated as reference category

| Effect | 1 dose vaccine |  | 2 doses vaccine |  |
| --- | --- | --- | --- | --- |
|  | Rate ratio (95% CI) | *p*-value | Rate ratio (95% CI) | *p*-value |
| No risk condition | 0.36 (0.33, 0.39) | $2 \times 10^{-118}$ | 0.14 (0.12, 0.15) | $2 \times 10^{-408}$ |
| Moderate risk condition | 0.41 (0.37, 0.45) | $2 \times 10^{-70}$ | 0.17 (0.15, 0.18) | $10 \times 10^{-304}$ |
| Eligible for shielding | 0.58 (0.50, 0.69) | $5 \times 10^{-11}$ | 0.32 (0.29, 0.37) | $1 \times 10^{-75}$ |

Presentation dates from 1 December 2020 to 8 September 2021.
Controls matched for age, sex, and primary care practice on date of presentation of case.
Adjusted for care home residence, number of adults in household, number of drug classes and recent hospital stay as covariates
Rate ratios estimated in multivariable conditional logistic regression with all variables in model
Vaccination status is doses administered at least 14 days before presentation date

**Table S3.** Rate ratios for hospitalised or fatal COVID-19 associated with vaccine dose within clinically extremely vulnerable subgroups, with unvaccinated as reference category

| Effect | 1 dose vaccine |  | 2 doses vaccine |  |
| --- | --- | --- | --- | --- |
|  | Rate ratio (95% CI) | *p*-value | Rate ratio (95% CI) | *p*-value |
| Solid organ transplant | 0.70 (0.33, 1.46) | 0.3 | 0.60 (0.38, 0.95) | 0.03 |
| Specific cancers | 0.88 (0.59, 1.32) | 0.5 | 0.31 (0.23, 0.41) | $2 \times 10^{-16}$ |
| Severe respiratory | 0.55 (0.43, 0.70) | $9 \times 10^{-7}$ | 0.28 (0.24, 0.33) | $3 \times 10^{-50}$ |
| Rare diseases | 0.48 (0.25, 0.92) | 0.03 | 0.31 (0.19, 0.50) | $1 \times 10^{-6}$ |
| On immunosuppressants | 0.74 (0.47, 1.15) | 0.2 | 0.40 (0.29, 0.54) | $2 \times 10^{-9}$ |
| Additional conditions | 0.46 (0.33, 0.63) | $2 \times 10^{-6}$ | 0.32 (0.26, 0.39) | $7 \times 10^{-26}$ |

Presentation dates from 1 December 2020 to 8 September 2021.
Severe COVID-19 defined as entry to critical care or death within 28 days of presentation.
Controls matched for age, sex, and primary care practice on date of presentation of case.
Adjusted for care home residence, number of adults in household, number of drug classes and recent hospital stay as covariates
Rate ratios estimated in multivariable conditional logistic regression with all variables in model
Vaccination status is number of doses administered at least 14 days before presentation date

**Table S4.** Rate ratios for hospitalized or fatal COVID-19 within risk groups associated with vaccine dose by product within risk groups, with unvaccinated as reference category

| Effect | 1 dose vaccine |  | 2 doses vaccine |  |
| --- | --- | --- | --- | --- |
|  | Rate ratio (95% CI) | *p*-value | Rate ratio (95% CI) | *p*-value |
| No risk condition: mRNA vaccine | 0.26 (0.23, 0.29) | $4 \times 10^{-101}$ | 0.08 (0.07, 0.09) | $4 \times 10^{-235}$ |
| No risk condition: AZ vaccine | 0.52 (0.46, 0.58) | $1 \times 10^{-29}$ | 0.19 (0.17, 0.20) | $9 \times 10^{-236}$ |
| Moderate risk condition: mRNA vaccine | 0.36 (0.31, 0.41) | $3 \times 10^{-47}$ | 0.09 (0.08, 0.11) | $5 \times 10^{-220}$ |
| Moderate risk condition: AZ vaccine | 0.48 (0.42, 0.54) | $6 \times 10^{-31}$ | 0.22 (0.20, 0.25) | $3 \times 10^{-186}$ |
| Eligible for shielding: mRNA vaccine | 0.41 (0.30, 0.55) | $2 \times 10^{-9}$ | 0.23 (0.19, 0.29) | $3 \times 10^{-41}$ |
| Eligible for shielding: AZ vaccine | 0.71 (0.59, 0.86) | $3 \times 10^{-4}$ | 0.38 (0.33, 0.43) | $7 \times 10^{-51}$ |

Presentation dates from 1 December 2020 to 8 September 2021.
Adjusted for care home residence, number of adults in household, number of drug classes dispensed, recent hospital stay and risk category.
Controls matched for age, sex, and primary care practice on date of presentation of case.
Vaccination status is number of doses administered at least 14 days before presentation date

**Table S5.** Risk factors for hospitalised or fatal COVID-19 in those who had received 2 doses of vaccine at least 14 days before

|  | Controls<br>(8225) | Cases (641) | Univariate |  | Multivariable |  |
| --- | --- | --- | --- | --- | --- | --- |
|  |  |  | Rate ratio (95%<br>CI) | *p*-value | Rate ratio (95%<br>CI) | *p*-value |
| Care home | 1127 (2%) | 213 (5%) | 1.90 (1.59, 2.26) | $7 \times 10^{-13}$ | 0.81 (0.66, 1.00) | 0.05 |
| SIMD quintile (integer) | 3 (1 to 4) | 2 (1 to 4) | 0.81 (0.78, 0.83) | $2 \times 10^{-49}$ | 0.88 (0.86, 0.91) | $5 \times 10^{-13}$ |
| Number of adults in household | 2 (1 to 2) | 2 (1 to 2) | 1.02 (0.99, 1.05) | 0.3 | 1.05 (1.02, 1.09) | 0.002 |
| Number of hospital diagnoses | 0 (0 to 3) | 4 (1 to 7) | 1.28 (1.27, 1.29) | $2 \times 10^{-448}$ | 1.09 (1.08, 1.11) | $5 \times 10^{-37}$ |
| Number of non-cardiovascular drug classes | 2 (1 to 5) | 6 (3 to 9) | 1.23 (1.21, 1.24) | $4 \times 10^{-446}$ | 1.11 (1.10, 1.12) | $2 \times 10^{-74}$ |
| Recent hospital stay | 925 (2%) | 830 (20%) | 11.6 (10.4, 13.1) | $9 \times 10^{-385}$ | 6.6 (5.8, 7.5) | $5 \times 10^{-185}$ |
| **Risk group** |  |  |  |  |  |  |
| No risk condition | 30516 (62%) | 999 (25%) |  | . |  | . |
| Moderate risk condition | 15102 (31%) | 1925 (47%) | 3.62 (3.32, 3.95) | $1 \times 10^{-184}$ | 1.80 (1.63, 1.99) | $2 \times 10^{-30}$ |
| Solid organ transplant | 97 (0%) | 94 (2%) | 29.9 (21.0, 42.7) | $2 \times 10^{-78}$ | 9.2 (6.2, 13.7) | $1 \times 10^{-27}$ |
| Specific cancers | 449 (1%) | 154 (4%) | 9.8 (8.0, 12.2) | $5 \times 10^{-100}$ | 3.86 (3.04, 4.90) | $1 \times 10^{-28}$ |
| Severe respiratory | 1599 (3%) | 454 (11%) | 8.3 (7.2, 9.5) | $3 \times 10^{-209}$ | 2.87 (2.45, 3.37) | $1 \times 10^{-38}$ |
| Rare diseases | 162 (0%) | 46 (1%) | 8.2 (5.7, 11.8) | $1 \times 10^{-30}$ | 3.46 (2.33, 5.15) | $8 \times 10^{-10}$ |
| On immunosuppressants | 461 (1%) | 117 (3%) | 8.3 (6.6, 10.5) | $1 \times 10^{-68}$ | 3.74 (2.87, 4.86) | $9 \times 10^{-23}$ |
| Additional conditions | 810 (2%) | 279 (7%) | 10.1 (8.6, 12.0) | $2 \times 10^{-163}$ | 3.67 (3.02, 4.46) | $6 \times 10^{-39}$ |

Percentages are column percentages for each variable
Rate ratios are from conditional logistic regression models matched for age, sex and general practice
Univariate rate ratios are for models with a single covariate
Multivariable rate ratios are for a model including all covariates shown in the table

**Table S6.** Incidence of severe COVID-19 in clinically extremely vulnerable individuals who had received 2 doses of vaccine at least 14 days before, by risk subgroup

| Risk group | Number of severe cases | Person-months | Rate / 1000 |
| --- | --- | --- | --- |
| Solid organ transplant | 35 | 23678 | 1.48 |
| Specific cancers | 33 | 93896 | 0.35 |
| Additional / rare conditions | 59 | 210801 | 0.28 |
| Severe respiratory | 80 | 303250 | 0.26 |
| On immunosuppressants | 30 | 118649 | 0.25 |

## Notes

### Competing Interest Statement

HC receives research support and honoraria and is a member of advisory panels or speaker bureaus for Sanofi Aventis, Regeneron, Novartis, Novo-Nordisk and Eli Lilly. HC receives or has recently received non-binding research support from AstraZeneca and Novo-Nordisk. SH received honoraria from Gilead.

### Funding Statement

No specific funding was received for this study. HC is supported by an endowed chair from the AXA Research Foundation.

### Author Declarations

This study was performed within Public Health Scotland as part of its statutory duty to monitor and investigate public health problems. Under the [UK Policy Framework for Health and Social Care Research](https://www.hra.nhs.uk/planning-and-improving-research/policies-standards-legislation/uk-policy-framework-health-social-care-research/) set out by the NHS Health Research Authority, this does not fall within the definition of research and ethical review was therefore not required. This has been confirmed in writing by the NHS West of Scotland Research Ethics Service. Individual consent is not required for Public Health Scotland staff to process personal data to perform specific tasks in the public interest that fall within its statutory role. The statutory basis for this is set out in Public Health Scotland's [privacy notice](https://www.publichealthscotland.scot/our-privacy-notice/personal-data-processing/).

### Summary of Updates

This version of the manuscript has been revised to include data extracted from health records up to 22 September 2021.

